# Protective effect of a first SARS-CoV-2 infection from reinfection: a matched retrospective cohort study using PCR testing data in England

**DOI:** 10.1101/2022.01.10.22268896

**Authors:** Joanne Lacy, Anna Mensah, Ruth Simmons, Nick Andrews, M. Ruby Siddiqui, Antoaneta Bukasa, Shennae O’Boyle, Helen Campbell, Kevin Brown

**Affiliations:** Immunisation and Vaccine Preventable Diseases Division, UK Health Security Agency, London, UK

## Abstract

The duration of immunity after first SARS-CoV-2 infection and the extent to which prior immunity prevents reinfection is uncertain and remains an important question within the context of new variants.

Using a retrospective population-based matched observational study approach, we identified cases with a first PCR positive test between 01 March 2020 and 30 September 2020 and cases were matched by age, sex, upper tier local authority of residence and testing route to individuals testing negative in the same week (controls) by PCR. After a 90-day pre-follow up period for cases and controls, any subsequent positive tests up to 31 December 2020 and deaths within 28 days of testing positive were identified, this encompassed an essentially vaccine-free period.

There were 517,870 individuals in the matched cohort with 2,815 reinfection cases and 12,098 first infections. The protective effect of a prior SARS-CoV-2 PCR-positive episode was 78% (OR 0.22, 0.21-0.23). Protection rose to 82% (OR 0.18, 0.17-0.19) after a sensitivity analysis excluded 934 individuals with a first test between March and May and a subsequent positive test between June and September 2020.

Amongst individuals testing positive by PCR during follow-up, reinfection cases had 77% lower odds of symptoms at the second episode (adjusted OR 0.23, 0.20-0.26) and 45% lower odds of dying in the 28 days after reinfection (adjusted OR 0.55, 0.42-0.71).

Prior SARS-CoV-2 infection offered protection against reinfection in this population. There was some evidence that reinfections increased with the Alpha variant compared to the wild-type SARS-CoV-2 variant highlighting the importance of continued monitoring as new variants emerge.

## Introduction

As of 10 December 2021, 267.9 million COVID-19 cases and 5.3 million associated deaths have been reported globally to the World Health Organisation(1). The first case of COVID-19 disease in the UK was confirmed in January 2020 and by the end of that same year, 52 million PCR tests had been performed with 2.65 million individuals testing positive at least once and 76,406 deaths reported within 28 days of a positive result. Reinfection is associated with seasonal human coronaviruses such as the common cold and there is a recognised need to understand the risk of reinfection in individuals that have recovered from COVID-; a population that will continue to increase.

The duration of immunity after first SARS-CoV-2 infection and the extent to which prior immunity provides protection against disease or transmission remains uncertain. Potential cases of reinfection have been reported internationally(2–6). In England, possible reinfection cases are defined as individuals that have an interval of at least 90 days between two consecutive positive tests. As of 31 October 2021, there have been 72,264 possible reinfections in England out of 7.8 million first positive tests. Confirmed reinfections are those with genetically distinct specimens at each episode, however these require sequencing at each episode so currently only a small proportion of possible reinfections can be confirmed. The most recent data up to 31 October 2021 included 441 confirmed reinfections(7).

An analysis of a large, multi-centre, prospective cohort study of hospital healthcare workers in the UK, found that a prior history of SARS-CoV-2 infection was associated with an 84% (95% CI: 81%-87%) lower risk of infection during 7 months of follow up(8). Similarly, a population-based study in Denmark found prior infection conferred 80% protection against reinfection in those aged 65 years and younger but only 47% protection against reinfection in those aged 65 years or older(9). A study of 43,000 individuals in Qatar estimated 95.2% (95% CI: 94.1-96.0%) protection against reinfection(10). A retrospective cohort study of patients in a health system in Ohio and Florida found that the protection offered from prior infection to reinfection was 81.8% (95% CI: 76.6-85.8) at ≥90 days after initial testing(11). The protection against symptomatic infection was 84.5% (95% CI: 77.9-89.1). A French study of SARS-CoV-2 screening tests found 42.6% of patients presented a similar clinical status upon reinfection (after ≥90 days), 29.5% had a milder form of the disease and 27.8% worsened from asymptomatic to mild or severe disease(12).

The emergence of the Alpha(13), Beta(14), Gamma(15), Delta(16) and most recently Omicron (17) SARS-CoV-2 variants was a cause for concern, not only for the apparent increased transmissibility of subsequent variants(13,18), associated with mutations in the spike protein(19,20) but also for the increased risk of hospitalisation(21) and the potential to escape the immune response(22–27). It is important to understand if both natural and vaccine-acquired immunity provides protection from SARS-CoV-2 variants.

Using national testing datasets in England we identified people who tested PCR positive for SARS-CoV-2 for the first time between 01 March 2020 and 30 September 2020, as cases. These individuals were matched one-to-one to controls with a negative test result in the same week of test and tested via the same testing route with the same demographic characteristics of age (in years), sex and upper tier local authority of residence. We then looked at the risk in both case and control groups of reporting a positive PCR test within a predetermined follow up period starting 90 days after the last COVID-19 PCR positive test of the cases up until 31 December 2020. This period therefore predated COVID-19 vaccine effects as vaccination began on 8 December 2020 in highest priority groups and anti-spike IgG levels mature over the 2-4 weeks after the first dose(28).

## Methods

### Study population and matching

The main analysis was a retrospective population-based matched observational study. COVID-19 is a notifiable disease and all laboratories in England are legally obliged to report all SARS-CoV-2 positive test results to Public Health England (PHE) and its successor, the UK Health Security Agency (UKHSA) through the Second Generation Surveillance System (SGSS)(29). The Unified Dataset (USD) consolidates data on positive tests from SGSS with national negative test data on an individual level. All SARS-CoV-2 testing undertaken by NHS or PHE laboratories for hospitalised patients or routine checks for healthcare workers are reported through pillar 1. Testing within the wider community for people reporting symptoms and requesting a test, regular testing of care home residents and their staff are reported through pillar 2.

Due to the limited number of paediatric reinfection cases identified in 2020, individuals in England aged 10 years or older were selected based on a first SARS-CoV-2 positive PCR test result between 01 March 2020 and 30 September 2020. This ensured that those selected with a positive test, and those matched with an initial negative PCR result, had at least 90 days to become a reinfection or first infection respectively before the end of the study period on 31 December 2020 and enabled review of reinfection risk prior to widespread introduction of vaccination in England. Upper tier local authority of residence was derived from the patient’s postcode or, if this was not available, then from their GP’s postcode. Individuals with missing postcodes or with the same postcodes as their testing laboratory were excluded.

To be eligible, cases could not test positive for a period of 90 days after the latest positive PCR test from their first COVID-19 episode. Anyone who remained PCR positive for longer than 60 days was excluded from analysis as a possible case of persistent infection. All individuals were linked with the Personal Demographics Service (PDS) of the NHS spine which stores demographic information including deaths for all NHS patients(30). We excluded all participants that died before the end of their assigned pre-follow-up 90-day period and those that could not be linked to PDS.

Two rounds of matching were carried out to match PCR-positive cases to individuals testing PCR negative in the same week with the same sex, age in years, pillar test route (community or hospital-based testing) and upper tier local authority [Figure 1]. The selection of controls from within the national testing dataset enabled identification of controls from the same population as the cases with assumed similar risk of infection to their matched cases. Individual matching was undertaken to directly compare the rates of infection while excluding potential confounders. Cases that had multiple positive tests after their first positive test for a period of less than 60 days were matched to an individual that tested negative based on first PCR test date but the study follow-up period for both the case and matched control started 90 days after the last positive test of the first episode of the case [Supplementary figure 1]. To be eligible, matched controls could not have tested positive before their selection, they could not have a positive result for the 90 day or full pre-follow up period required by their matched case. Matched controls were also linked to PDS and were excluded if they died during the pre-follow up period or could not be found on PDS.

**Figure 1.**
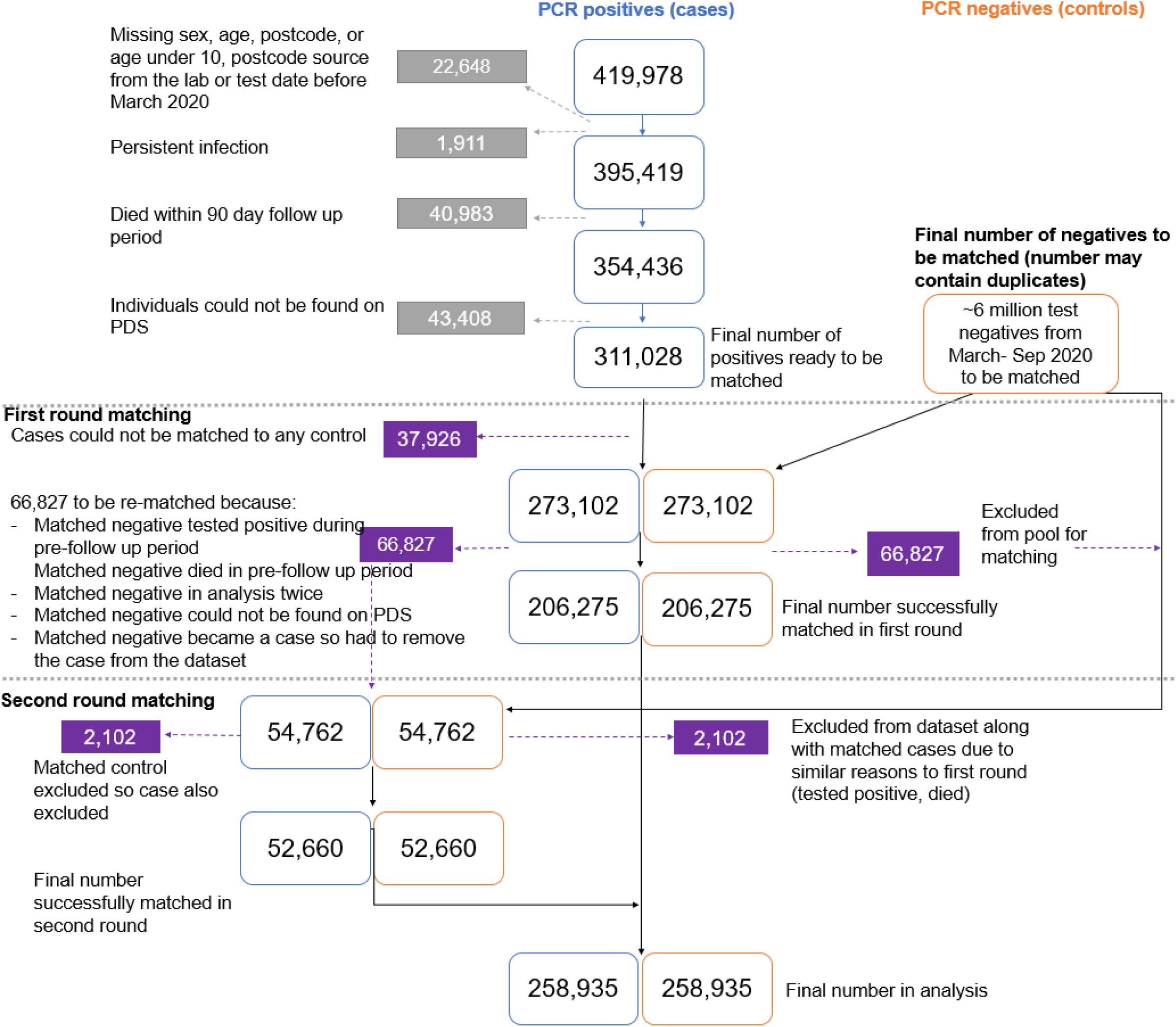
Diagram detailing the cleaning and matching steps taken to select the final study population

We matched on a month by month basis to exclude matched negatives from becoming part of the study for a second time if they tested negative again after they had already been linked to a case from an earlier month. Both the cases and controls were included if they had previous negative tests prior to inclusion in the study, however neither could have had a prior positive test. Additional details on both cases and controls were extracted including whether they had indicated symptoms at their first test, their ethnicity which was obtained from the National Immunisation Management System (NIMS) and their Indices of Multiple Deprivation (IMD) quintile which was obtained from the individuals’ postcode and linked to the 2019 indices of deprivation data; an IMD quintile of 1 represents individuals with postcodes in the most deprived areas.

### Follow-up period

The follow up period started after the 90-day pre-follow up period was completed and continued until 31 December 2020. The start of the follow up period varied depending on when the case first tested positive. Cases that first tested positive on 1 March 2020 had an earliest start date of 30 May 2020 for example, while some cases that first tested positive in September 2020 had their follow up period start in December 2020.

The cases and controls were linked by NHS number and via their unique identifier assigned by SGSS to all records of positive SARS-CoV-2 test results to find any subsequent PCR-positive test results. We used lookup tables to match the controls’ unique identifier from the USD to their unique identifier in SGSS. Reinfection cases were identified from the main SGSS dataset through identification of 90-day or longer intervals between sequential positive results for the same individual and this data was linked to our case data via NHS number and unique identifier.

We extracted extra information from PCR tests in the follow up period including whether it was community or hospital-based testing and, for those individuals that were tested through community testing, we also obtained information on whether the participant was symptomatic together with data on the PCR results for the target genes. Three of the four Lighthouse laboratories used a diagnostic assay that does not detect a deletion of amino acids 69 and 70 of the spike protein, also known as S gene target failure (SGTF). By 15 November 2020, S gene target failure became a reliable proxy for the Alpha (Kent B.1.1.7) variant that emerged in the UK in September and was associated with increased infectivity compared to the wild-type SARS-CoV-2 variant. We identified deaths up to 60 days beyond the end of the follow up period using PDS records.

Controls that subsequently tested positive were excluded from the case population.

### Statistical analysis

We used descriptive analysis to describe the population testing positive during the follow up period up. We carried out logistic regression to give the univariable odds ratios for testing positive for each of the matching variables, ethnicity and IMD quintile. The upper tier local authorities were grouped into the nine UKHSA regions for the analysis [Table 1].

**Table 1.**
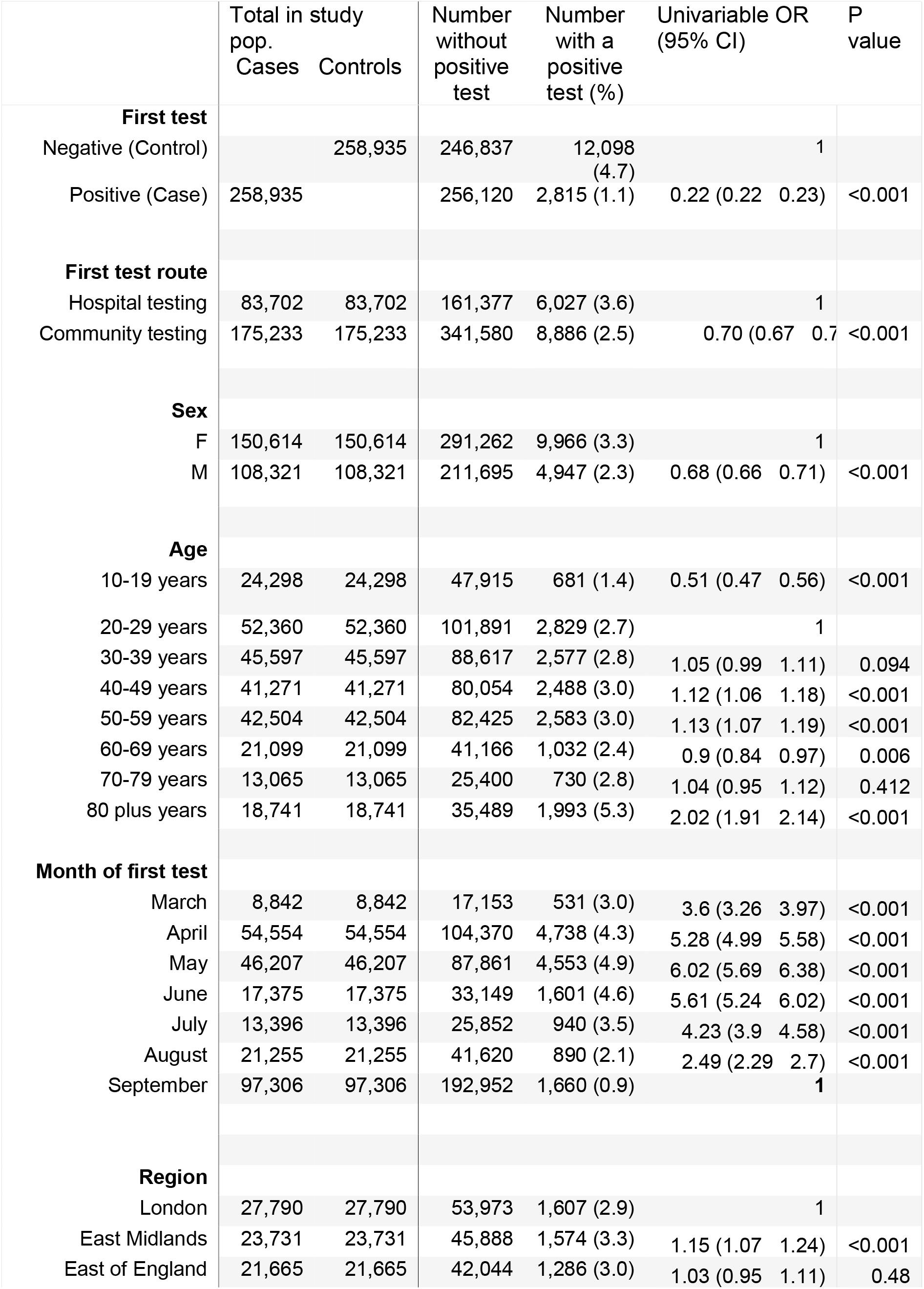
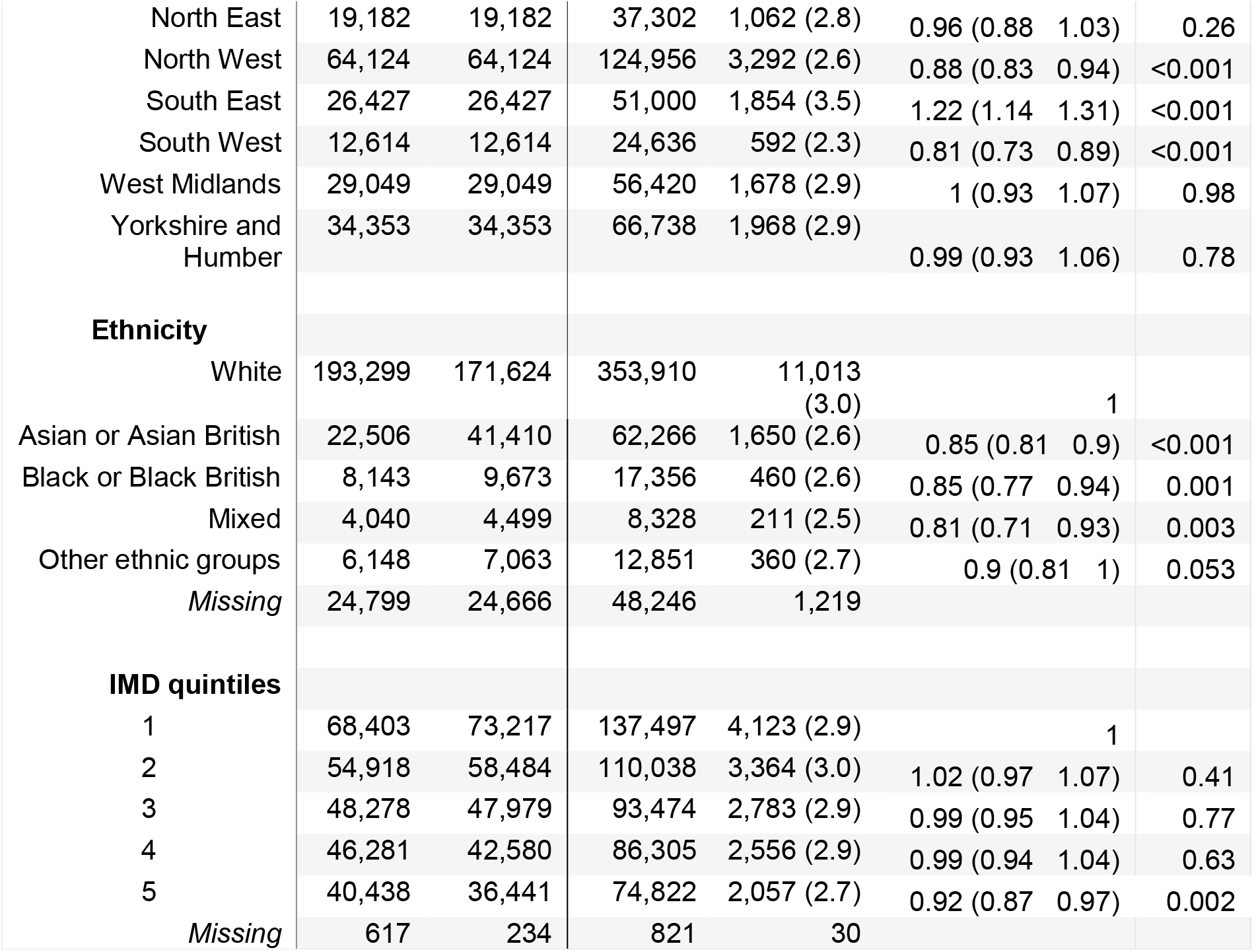
Description of study population in terms of matching variables (first test pillar, sex, age, month of first test, region), ethnicity, IMD quintiles and distribution of the outcome, tested positive, with univariable odds ratios from logistic regression

Conditional logistic regression was used to take into account the matching design and to give an adjusted odds ratio for the cases and controls testing positive during the follow up period. We added predictors that were not matched (ethnicity, IMD quintile) into the conditional model to test for confounding. As the conditional regression model already accounted for the matched variables, these were not included in the final model.

A sensitivity analysis was done by splitting the follow up time into 30-day groupings to compare the odds of testing positive in the cases compared to the controls during each 30 days of follow up while excluding those that died or tested positive before the start of each time point. We carried out conditional logistic regression on each 30-day time point while stratifying by month to identify all time-related effects.

Secondary outcomes within the population that tested positive during the follow-up period were also monitored. We used logistic regression to compare the odds of dying within 28 days, odds of dying within 60 days, odds of symptomatic infection and odds of second test being S gene-negative between cases and controls. We used logistic regression for this analysis instead of conditional logistic regression due to the small numbers of matched pairs where each of the matched pair tested positive. Univariable odds ratios were calculated for each predictor and then a multivariable model was built. We tested additional variables one by one in the model with the likelihood ratio test. Age categories were regrouped into a binary variable with those aged over 50 and those aged 10-49 due to small numbers. Region was re-categorized for the S gene analysis into one region with London, South East and East of England and the other category with the rest of England. The first of these regional groupings roughly corresponded to the regions with Alpha variant dominance in November and December 2020.

### Post-hoc analysis

After the sensitivity analysis, the dataset was trimmed for a post-hoc conditional logistic regression analysis which excluded 2,987 cases and their matched controls that had their first test in either March, April or May 2020 and then had a positive test or died in either the first 90 days (individuals from March), first 60 days (individuals from April) or first 30 days (Individuals from May) of follow up. This corresponds to excluding 934 individuals that tested positive in the summer months between June and September 2020; these cases and controls were excluded due to the stratified results of the conditional regression of the whole dataset and concerns about the quality and limited testing during the early months of the pandemic [Figure 2].

**Figure 2.**
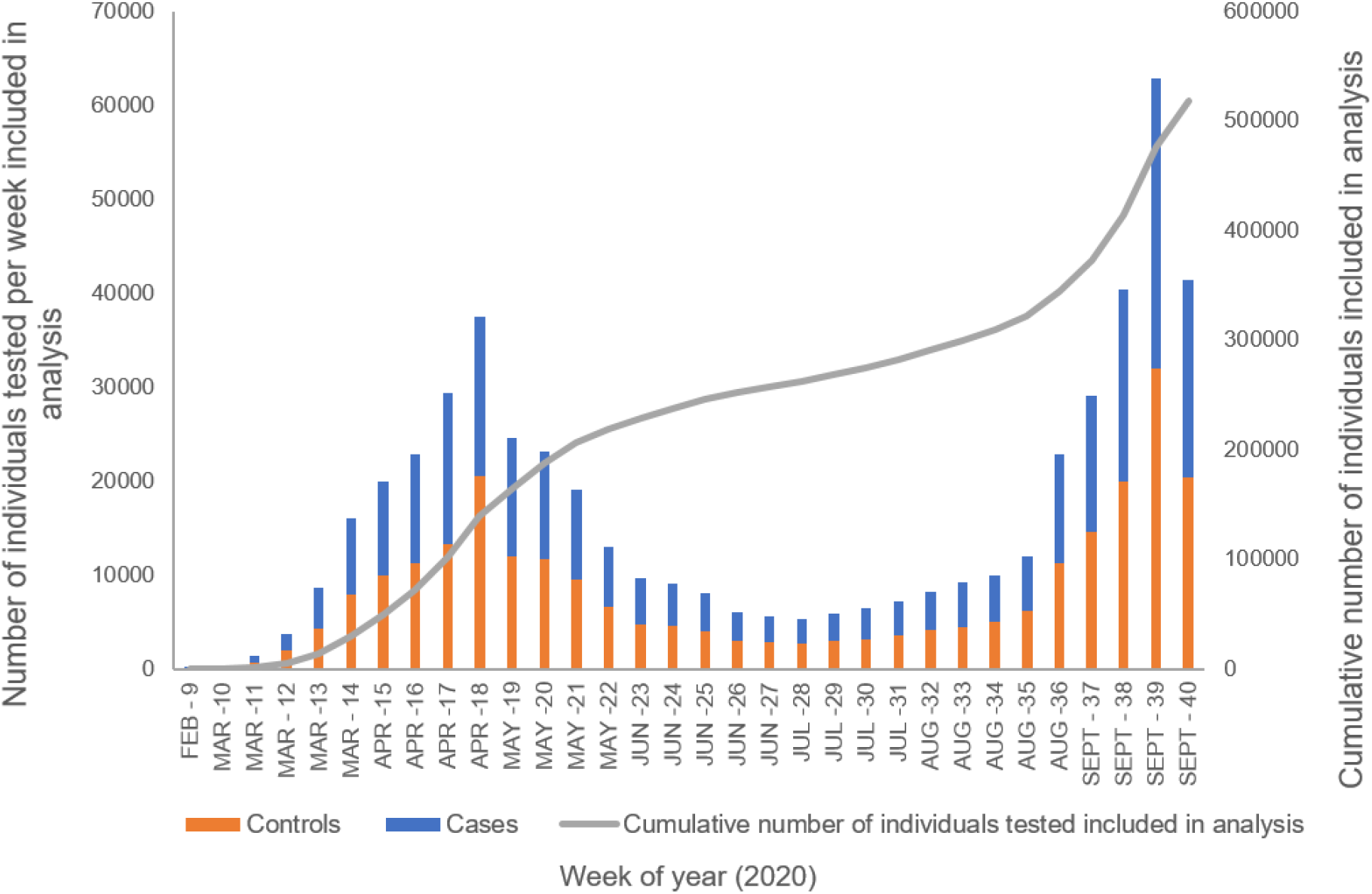
Distribution by week of test for total individuals (cases and controls) included in final analysis

Statistical analysis was carried out in STATA version 15.1, linkage to SGSS and USD datasets was carried out in Microsoft SQL server management studio 18.

## Results

### Study population and matching

There were 419,978 individuals that tested positive by PCR for SARS-CoV-2 between March and September 2020 in England. We excluded 22,648 individuals that either had missing demographic information or were under 10 years old, 1,911 individuals with persistent infection, 40,983 individuals that died in the pre-follow up period and 43,408 individuals that could not be found on PDS. The remaining 311,028 individuals with positive PCR tests underwent two rounds of matching to a test-negative with the same sex, age, week of test, upper tier local authority of residence and pillar of testing. The first round of matching led to 66,827 exclusions of the matched controls due to the controls either testing positive in the pre-follow up period, dying within 90 days of their first test, not linking in PDS or being included in the study twice and therefore a second matching round was undertaken for these individuals [Figure 1]. There were 638 negative controls that became positive cases and so these individuals were kept in the control population but were removed from the case population along with their matched control.

The final cohort consisted of 258,935 successfully matched cases giving a total of 517,870 individuals.

### Descriptive analysis

Figure 2 shows the distribution by week of the 517,870 individuals included in the analysis by date of first test. There was a peak in April 2020 and then a lull in the summer months with a subsequent increase in positive tests from September 2020. Data for the last week of September is incomplete as 30 September 2020 fell on a Wednesday and so there was only data for three days instead of seven.

During the follow up period 14,913 individuals had a positive test; 2,815 were reinfections in the cases and the remaining 12,098 positive tests were first infections in the controls. Cases had 78% lower odds of testing positive in the follow up period than controls (univariable odds ratio 0.22, 95% CI 0.22-0.23). There was a peak in positive tests in the controls at the beginning of the follow up period and subsequent increase in positive tests around 100 days of follow up [Figure 3]. The distribution of test-positives in cases was more linear and remained lower than in controls. A decreasing number of individuals contributed to the analysis over time.

**Figure 3.**
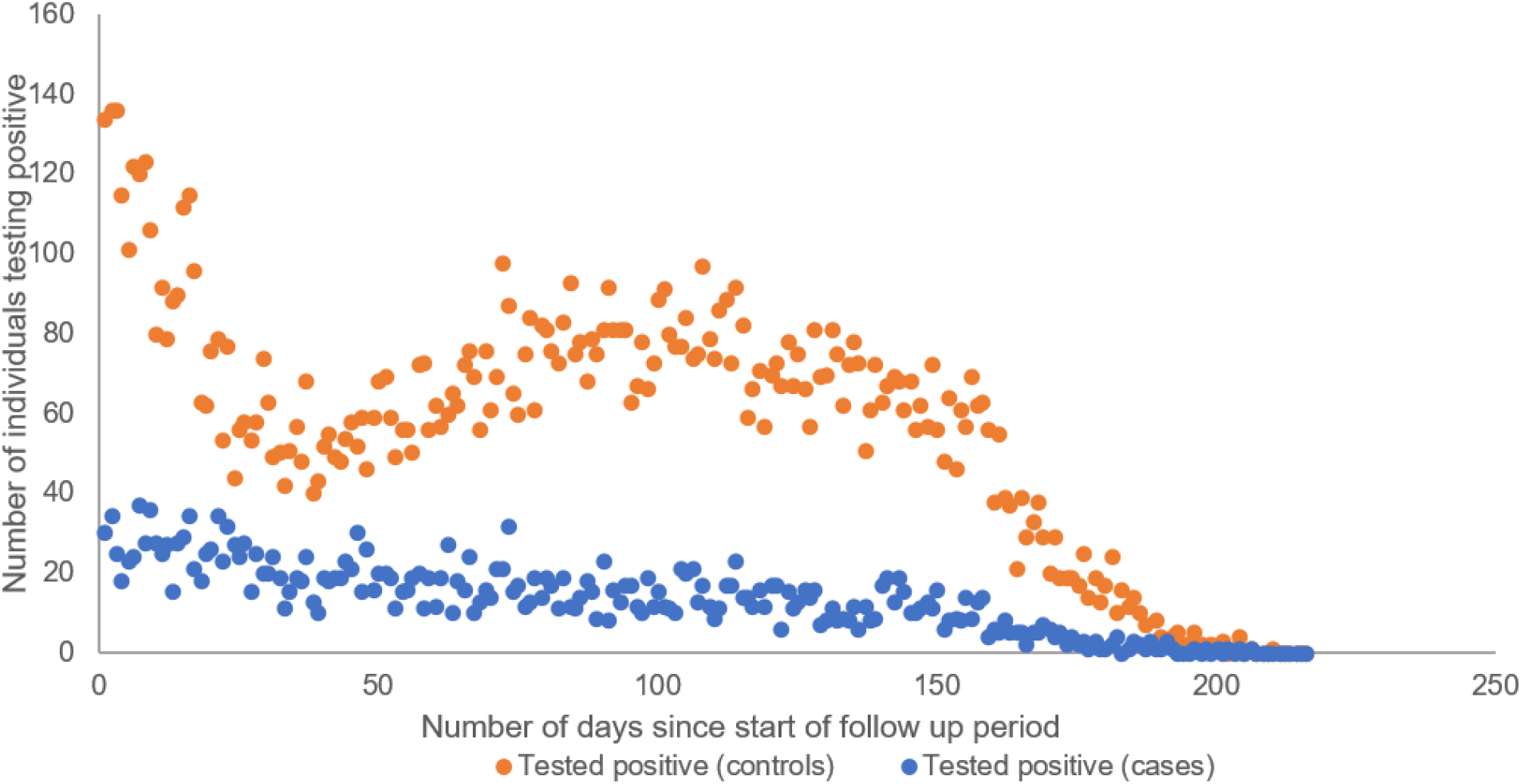
Distribution of positive tests during follow up period by days of follow up for cases and controls. The follow up period does not include the pre-follow up 90 day period after first tests

Based on the univariable analysis, men had 32% decreased odds (OR 0.68, 0.66-0.71) of testing positive in the follow-up period compared to women [Table 1], while those initially tested using community testing were also less likely to test positive in the follow up period (OR 0.70, 0.67-0.72) compared to those that had their first test in hospital-based testing. Children aged 10-19 had 49% lower odds of testing positive (OR 0.51, 0.47-0.56) compared to adults aged 20-29, while adults aged 80+ were twice as likely to have a positive test (OR 2.02, 1.91-2.14). Individuals with Asian or Asian British (OR 0.85, 0.81-0.9), Black or Black British (OR 0.85, 0.77-0.94) and Mixed (OR 0.81,0.71-0.93) ethnicities had lower odds of testing positive compared to individuals with white ethnicities.

The results of the overall conditional logistic regression which takes into account the matching variables, found cases had 78% lower odds (OR 0.22, 0.21-0.23) of a positive test during the follow up period compared to controls [Table 2]. As the variables used for matching were taken into account by the model, they do not give an output within the conditional logistic regression model and so are not included in Table 2. Ethnicity and IMD quintile were not added to the multivariable conditional logistic regression model as they were not found to be significant confounders.

**Table 2.** Results from the conditional logistic regression with univariable odds ratios from the full dataset and from the post-hoc conditional logistic regression analysis. Matched variables are not included in this table as there is no output when included in a conditional model.

|  |  |  |  |  | Post hoc analysis with trimmed dataset |  |  |  |
| --- | --- | --- | --- | --- | --- | --- | --- | --- |
|  | *Did not test positive* | *Tested positive* | *Univariable OR conditional model (95% CI)* | *P value* | *Did not test positive* | *Tested positive* | *Univariable OR conditional model (95% CI)* | *P value* |
| **First test** |  |  |  |  |  |  |  |  |
| Negative (Control) | 246,837 | 12,098 | 1 |  | 244,316 | 11,632 | 1 |  |
| Positive (Case) | 256,120 | 2,815 | 0.22 (0.21 0.23) | <0.001 | 253,708 | 2,240 | 0.18 (0.17 0.19) | <0.001 |

Once the dataset was split into different timepoints for a sensitivity analysis, viewing the odds ratios by month of first test revealed that the odds ratios vary by month of first test and follow-up time [Figure 4] with odds ratios as high as 3.25 (1.06-9.97) in individuals that tested positive during the first 30 days of follow-up with their initial test from March. The higher odds ratios for individuals with first tests in March, April and May during the beginning of follow up corresponds to higher odds of testing positive for the cases than controls during the months of June, July, August and September.

**Figure 4.**
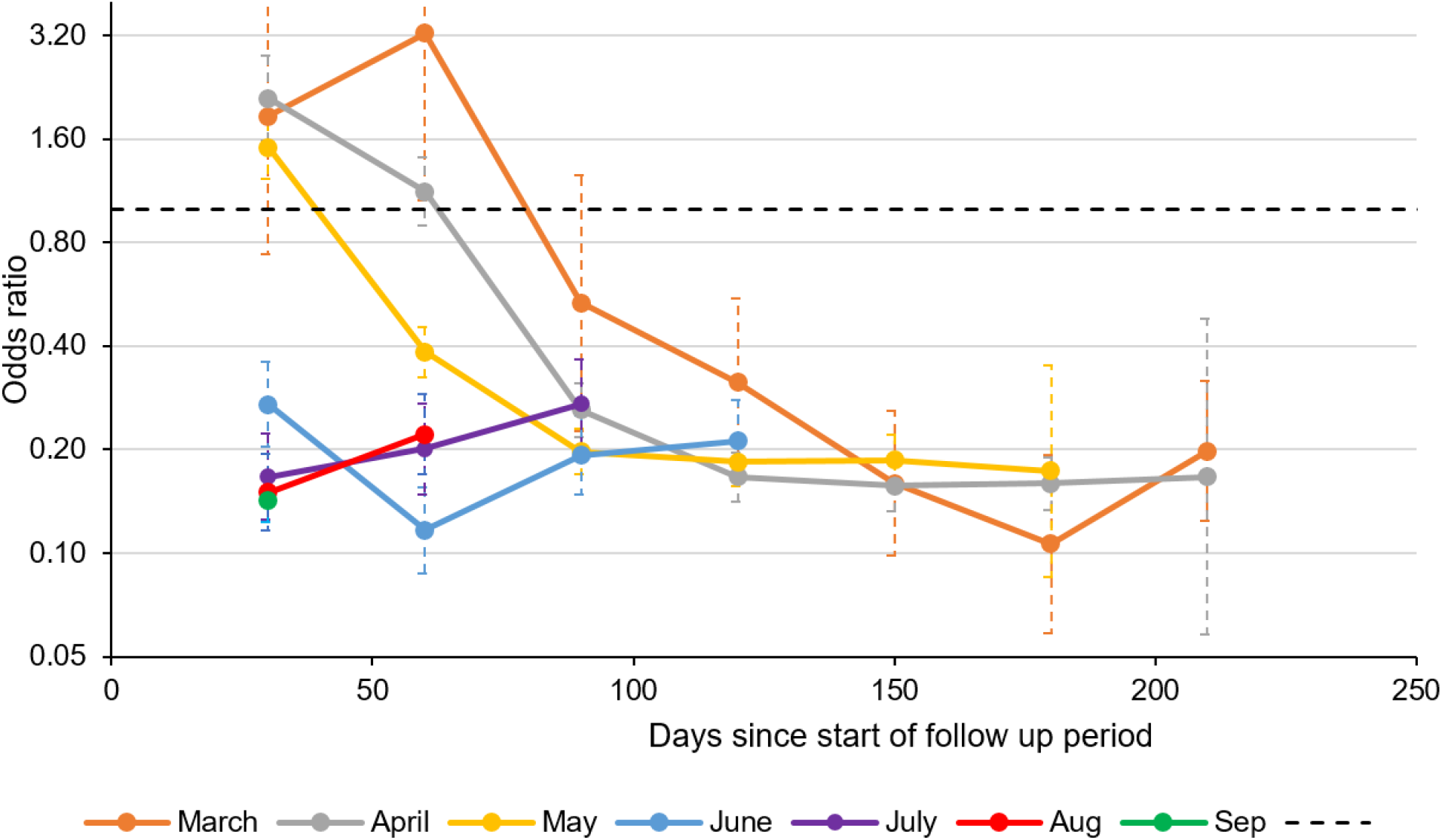
Adjusted odds ratios of a reinfection compared to a first infection according to month of original positive or negative test

The post-hoc analysis excluded 934 individuals that tested positive and 2,111 individuals that died. Those excluded had a first test during the months of March, April and May and had an event during the 90-day, 60 day and 30-day time point respectively. After additionally excluding the matched case or control for these individuals, the final post-hoc dataset consisted of 511,896 individuals with 255,948 individuals in each matched cohort. After excluding these individuals, the odds of testing positive during the follow up period for the cases was 82% (OR 0.18, 0.17-0.19) lower in cases than for controls.

### Secondary outcomes

There were 3,376 deaths amongst controls and 3,031 deaths amongst cases in those that did not have a PCR-positive COVID test during the follow up period. Of the 13,979 individuals in the post-hoc analysis that tested positive during the follow up period, 608 controls (5.2%) and 89 cases (3.9%) died within 28 days of the positive test and 703 (6.0%) controls and 111 (4.9%) of cases died within 60 days of the positive test [Table 3]. Individuals that were aged over 50 had more than 57 times the odds of dying compared to individuals aged 10-49 (OR 57.5, 36.40-90.84) and men had twice the odds of dying compared to women (OR 2.07, 1.78 -2.41).

**Table 3.**
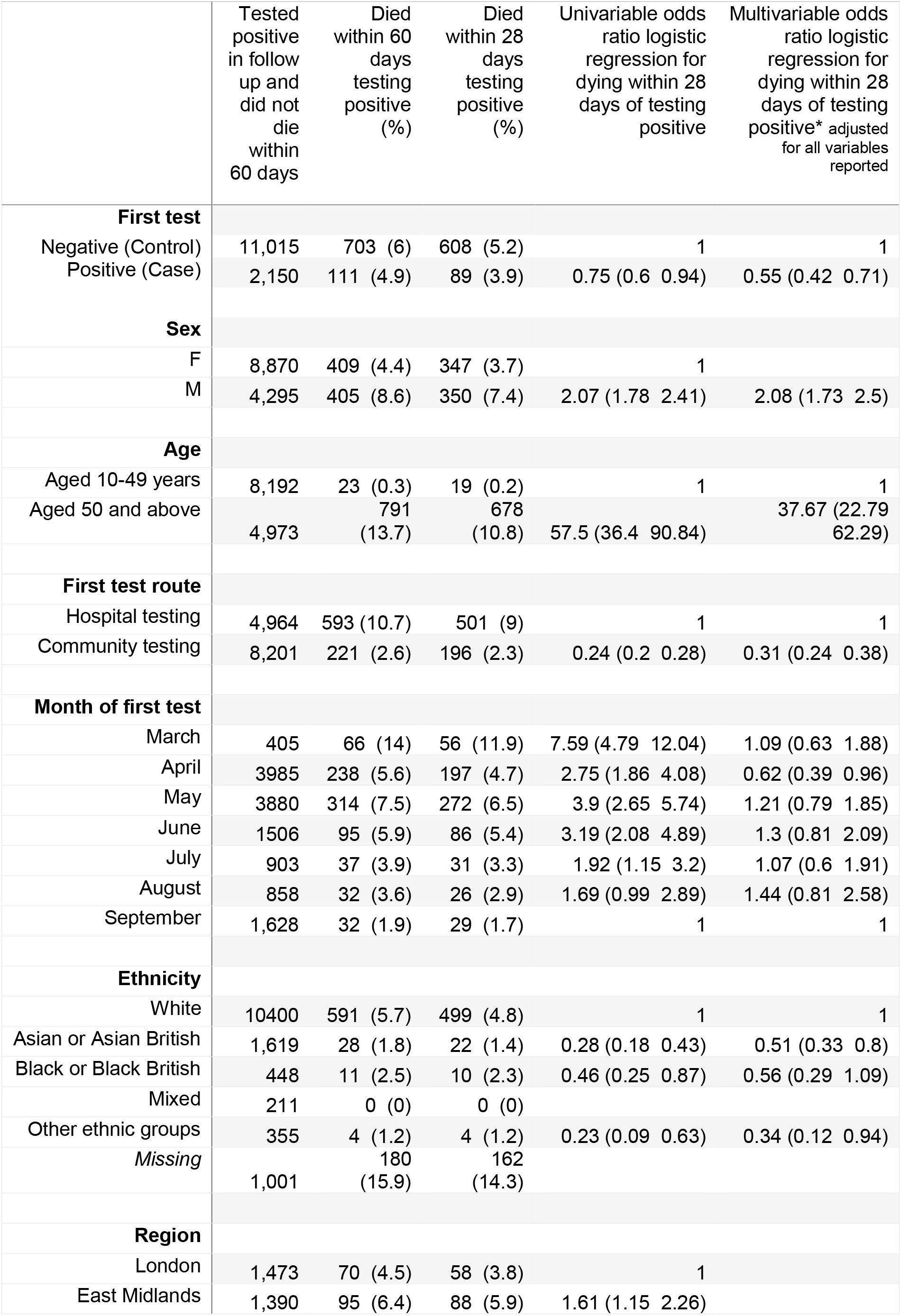
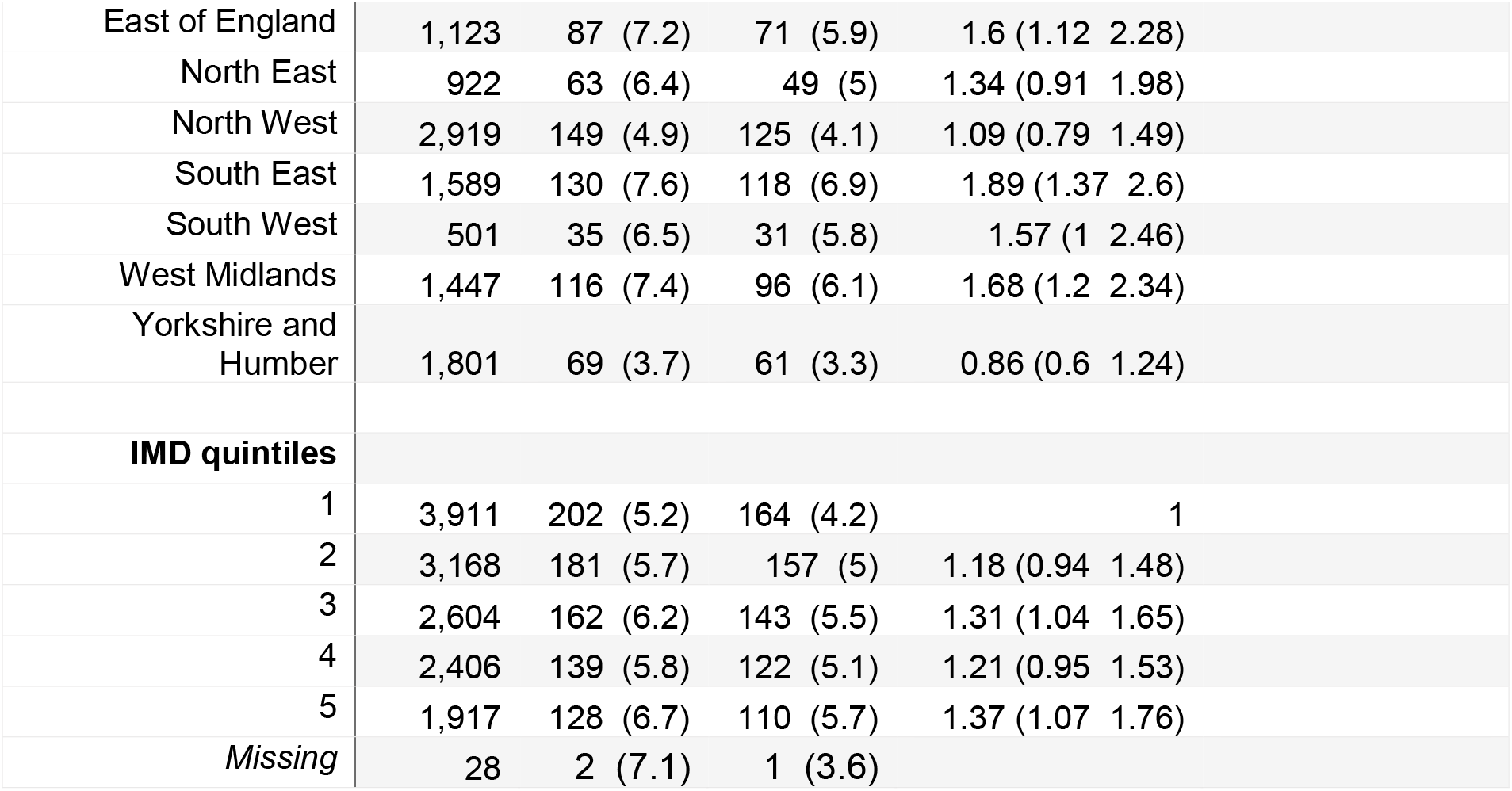
Deaths within those that tested positive for COVID during the follow up period from post hoc dataset along with results from logistic regression for odds of dying at 28 days in cases compared to controls

|  | Tested positive in follow up and did not die within 60 days | Died within 60 days testing positive (%) | Died within 28 days testing positive (%) | Univariable odds ratio logistic regression for dying within 28 days of testing positive | Multivariable odds ratio logistic regression for dying within 28 days of testing positive* adjusted for all variables reported |
| --- | --- | --- | --- | --- | --- |
| **First test** |  |  |  |  |  |
| Negative (Control) | 11,015 | 703 (6) | 608 (5.2) | 1 | 1 |
| Positive (Case) | 2,150 | 111 (4.9) | 89 (3.9) | 0.75 (0.6 0.94) | 0.55 (0.42 0.71) |
| **Sex** |  |  |  |  |  |
| F | 8,870 | 409 (4.4) | 347 (3.7) | 1 |  |
| M | 4,295 | 405 (8.6) | 350 (7.4) | 2.07 (1.78 2.41) | 2.08 (1.73 2.5) |
| **Age** |  |  |  |  |  |
| Aged 10-49 years | 8,192 | 23 (0.3) | 19 (0.2) | 1 | 1 |
| Aged 50 and above | 4,973 | 791 (13.7) | 678 (10.8) | 57.5 (36.4 90.84) | 37.67 (22.79 62.29) |
| **First test route** |  |  |  |  |  |
| Hospital testing | 4,964 | 593 (10.7) | 501 (9) | 1 | 1 |
| Community testing | 8,201 | 221 (2.6) | 196 (2.3) | 0.24 (0.2 0.28) | 0.31 (0.24 0.38) |
| **Month of first test** |  |  |  |  |  |
| March | 405 | 66 (14) | 56 (11.9) | 7.59 (4.79 12.04) | 1.09 (0.63 1.88) |
| April | 3985 | 238 (5.6) | 197 (4.7) | 2.75 (1.86 4.08) | 0.62 (0.39 0.96) |
| May | 3880 | 314 (7.5) | 272 (6.5) | 3.9 (2.65 5.74) | 1.21 (0.79 1.85) |
| June | 1506 | 95 (5.9) | 86 (5.4) | 3.19 (2.08 4.89) | 1.3 (0.81 2.09) |
| July | 903 | 37 (3.9) | 31 (3.3) | 1.92 (1.15 3.2) | 1.07 (0.6 1.91) |
| August | 858 | 32 (3.6) | 26 (2.9) | 1.69 (0.99 2.89) | 1.44 (0.81 2.58) |
| September | 1,628 | 32 (1.9) | 29 (1.7) | 1 | 1 |
| **Ethnicity** |  |  |  |  |  |
| White | 10400 | 591 (5.7) | 499 (4.8) | 1 | 1 |
| Asian or Asian British | 1,619 | 28 (1.8) | 22 (1.4) | 0.28 (0.18 0.43) | 0.51 (0.33 0.8) |
| Black or Black British | 448 | 11 (2.5) | 10 (2.3) | 0.46 (0.25 0.87) | 0.56 (0.29 1.09) |
| Mixed | 211 | 0 (0) | 0 (0) |  |  |
| Other ethnic groups | 355 | 4 (1.2) | 4 (1.2) | 0.23 (0.09 0.63) | 0.34 (0.12 0.94) |
| Missing | 1,001 | 180 (15.9) | 162 (14.3) |  |  |
| **Region** |  |  |  |  |  |
| London | 1,473 | 70 (4.5) | 58 (3.8) | 1 |  |
| East Midlands | 1,390 | 95 (6.4) | 88 (5.9) | 1.61 (1.15 2.26) |  |

|  |  |  |  |  |
| --- | --- | --- | --- | --- |
| East of England | 1,123 | 87 (7.2) | 71 (5.9) | 1.6 (1.12 2.28) |
| North East | 922 | 63 (6.4) | 49 (5) | 1.34 (0.91 1.98) |
| North West | 2,919 | 149 (4.9) | 125 (4.1) | 1.09 (0.79 1.49) |
| South East | 1,589 | 130 (7.6) | 118 (6.9) | 1.89 (1.37 2.6) |
| South West | 501 | 35 (6.5) | 31 (5.8) | 1.57 (1 2.46) |
| West Midlands | 1,447 | 116 (7.4) | 96 (6.1) | 1.68 (1.2 2.34) |
| Yorkshire and Humber | 1,801 | 69 (3.7) | 61 (3.3) | 0.86 (0.6 1.24) |
| **IMD quintiles** |  |  |  |  |
| 1 | 3,911 | 202 (5.2) | 164 (4.2) | 1 |
| 2 | 3,168 | 181 (5.7) | 157 (5) | 1.18 (0.94 1.48) |
| 3 | 2,604 | 162 (6.2) | 143 (5.5) | 1.31 (1.04 1.65) |
| 4 | 2,406 | 139 (5.8) | 122 (5.1) | 1.21 (0.95 1.53) |
| 5 | 1,917 | 128 (6.7) | 110 (5.7) | 1.37 (1.07 1.76) |
| Missing | 28 | 2 (7.1) | 1 (3.6) |  |

The testing route of the first test was associated with dying as individuals tested through community testing had 76% lower odds of dying after a positive test compared to individuals that had their first test in hospital (OR 0.24, 0.20-0.28). After adjusting for sex, age, route of first test, month of first test and ethnicity, the cases went from having 25% lower odds of dying (OR 0.75, 0.60-0.94) to 45% lower odds of dying within 28 days of their reinfection compared to the odds of controls dying after their first infection (adjusted OR 0.55, 0.42-0.71).

Of those individuals in the post-hoc dataset that tested positive during the follow up period, symptom data was recorded for 75% of individuals, and the percentage of missing symptom data varied by age, month of first test, region and first test route [Supplementary table 1]. All tests with symptom data would have all been taken through the community testing route. Of controls that tested positive, 5,860 (66.3%) had symptoms, this compares to 499 (30.0%) of the reinfection cases. Men were more likely to report symptoms on their positive test with 1.8 times the odds of having symptoms at their second test compared to women (OR 1.75, 1.61-1.91), and all regions of England had lower odds of having symptoms compared to individuals from London. Individuals with Asian or British Asian ethnicity had twice the odds as individuals with White ethnicities to report symptoms (OR 2.12, 1.85-2.43) while individuals with Black or Black British ethnicities had 38% lower odds of reporting symptoms (OR 0.62, 0.50-0.77). After adjusting for sex, age group, month of first test, region and ethnicity, the odds of cases having symptoms during their reinfection was 77% (adjusted OR 0.23, 0.20-0.26) lower than for controls having symptoms during their first test.

The S-gene target failure (SGTF) was used as a proxy for individuals infected with the Alpha variant from 15 November 2020. There were 8,435 individuals that tested positive from 15 November, of these 37% had S-gene data available, the percentage of missing S-gene data varied by first test type, region, age, month of first test, first test route and ethnicity [Supplementary table 2]. There were 159 (54.6%) reinfection cases and 1,205 (42.5%) controls with SGTF. Individuals that lived outside of London, the South East and East of England had 84% lower odds of having SGTF (OR 0.16, 0.13-0.19). Older adults aged above 50 years old had 19% lower odds of SGTF (OR 0.81, 0.69 -0.95). After adjusting for region, age group, month of first test and ethnicity, reinfection cases had 1.9 times the odds of having SGTF (indicative of the Alpha variant) compared to controls with their first test (adjusted OR 1.90, 1.43-2.51).

## Discussion

Among our matched cohort of 517,870 individuals, there were 2,815 reinfection cases and 12,098 first infection cases up until 31 December 2020. Therefore, individuals with a prior SARS-CoV-2 PCR positive test had 78% lower odds (OR 0.22, 0.21-0.23) of having a second infection compared to individuals with no prior positive, from at least 90 days after their positive test up to a maximum of 9 months later. Protection increased to 82% lower odds (OR 0.18, 0.17-0.19) in our post-hoc analysis. These findings demonstrate a baseline of protection from a prior SARS-CoV-2 infection against reinfection before the widespread introduction of vaccination against COVID-19 in December 2020. This finding is in line with the results of a Danish study over a similar timeframe which found 80.5% protection against a second infection among 525,339 individuals that were followed in the second wave of the pandemic(9). Our findings are consistent with the SIREN study which followed healthcare workers in the UK and found that a previous infection was associated with a 84% lower risk of infection(8). The SIREN study was based on a similar period with data extracted up to January 2021, however the health care worker population were regularly tested and probably more likely to be exposed to COVID-19 than the general population. Our findings show that the protective effect of previous infection in the general population is at least very similar to that in health care workers, though we could be underestimating the protection as our study relies on individuals seeking COVID-19 testing and we postulate that seeking testing is a health seeking behaviour that could change after previous PCR confirmed SARS-CoV-2 infection. In those with a PCR-positive test during the follow-up period, the reinfection cases appeared to have milder disease with 77% lower odds of having symptoms at their second test (adjusted OR 0.23, 0.20-0.26). Age was significantly associated with death in all infections in the follow-up period with adults aged above 50 having 57 times the odds of dying (OR 57.5, 36.40-90.84) compared to younger adults and children aged 10-49 in the 28 days after testing positive. Cases of reinfection had 45% lower odds of dying in the 28 days after their reinfection compared to the controls (adjusted OR 0.55, 0.42-0.71).

The sensitivity analysis further investigated the odds of testing positive at different time points by month of first test thus revealing that there were higher odds of testing positive in the cases compared to the controls within individuals that had their first test in March, April and May and then had a positive test during the follow up period in June, July and August and September. The reasons for this disparity and high number of reinfections compared to the overall protective effect of prior SARS-CoV-2 infection are uncertain, however, it is possible that the population who were tested during the beginning of the pandemic were fundamentally different and were more likely to be elderly, hospitalised patients due to limited testing. Indeed 50.1% of individuals with a first test in March, April or May were aged over 50 years compared to 27.1% of individuals with a first test in June-September and 59.1% of individuals tested in these early months were tested through hospital testing compared to only 12.6% of individuals with first test in June-September. The uncertain reinfection cases occurred soon after the end of the 90-day pre-follow up period and so this could mean that some are persistent infections in this specific population. Cycle threshold (Ct) values are obtained from a PCR test and can give an indication of how much viral genetic material there is in a sample, a lower Ct value indicates that there is a higher concentration of virus, higher Ct values mean there is less virus and could indicate a persistent infection. However, Ct values are not recorded for all COVID-19 PCR tests in England and Ct values cannot be compared between different assays(31). As we do not adjust for comorbidities or look at Ct values in our analysis it is difficult to examine our hypothesis of persistent infections in detail, therefore we excluded 934 positive infections in cases and controls from the summer in the post-hoc analysis. It is also possible that a small number of positive PCR results in that period could be false positives due to the low prevalence of COVID-19 at that time and resultant lower positive predictive value (PPV) of testing leading to non-differential misclassification of the outcome in cases and controls.

Our analysis focuses on the 90-day criteria for *possible* reinfection and information is not available on the possible reinfection cases in our study to allow them to meet the *confirmed* reinfection definition. Large longitudinal cohort studies with epidemiological and virological information from each infection episode would be needed to differentiate cases that are SARS-CoV-2 RNA-positive over long periods of time from true cases of reinfection. It is also possible that individuals within the study could have had a reinfection over a shorter period than 90 days, the CDC also uses 45-day criteria for persons with COVID-19 like symptoms in addition to the 90 day interval criteria for individuals without symptoms, a 45 day interval was not investigated in our study due to incomplete symptom data(32).

Our results demonstrate that reinfection cases had almost twice the odds of being infected with the Alpha variant (adjusted OR 1.90, 1.43-2.51) compared to the controls, although these results should be interpreted with caution as we used S gene target failure as a proxy for the Alpha variant and there was a high proportion of missing S gene target failure data. The Delta variant emerged in spring 2021 and contributed to over 60% of sequenced cases in England by 17 May 2021(33). This was after the period covered by our analysis which focused on reinfections prior to the introduction of vaccines so that vaccination did not confound the results.

However, a UKHSA analysis found that the odds ratio of possible reinfection was higher with the Delta compared to the Alpha variant (OR 1.46, 1.03-2.05)(34), thus emphasising the need to monitor any increases in possible reinfection cases linked to a new variant. The emergence of Omicron highlights these concerns as analysis of routine surveillance data of almost 3 million individuals with a laboratory confirmed SARS-CoV-2 result from South Africa found that there was a greater risk of re-infection compared to primary infection during the current Omicron wave (hazard ratio 2.39, 95% CI 1.88–3.11) highlighting that Omicron can evade immunity, the study did not find evidence of increased reinfection for either the Beta and Delta variants (27).

As COVID-19 vaccine roll-out continues and some countries have started delivering booster programmes, it has become increasingly important to understand the impact of previous infection on the efficacy of COVID-19 vaccines and the need for boosting in a two-dose regimen. The COVID-19 vaccines currently in use trigger immune responses to the spike protein, however natural infection generates a broader humoral and cellular immune response with convalescent individuals demonstrating strong CD8+ T cell responses(35). The data on immune responses following Pfizer-BioNTech COVID-19 vaccination (BNT162b2) indicates that individuals with previous SARS-CoV-2 infection may generate stronger immune responses with more cross neutralising antibodies to one dose of BNT162b2 vaccine than compared to those without previous infection(36–39). There are concerns about waning protection post-vaccination against the Delta variant with one study showing protection against symptomatic disease reduced to 47.3 (95% CI 45 to 49.6) and 69.7 (95% CI 68.7 to 70.5) for Vaxzevria and BNT162b2, respectively after 20 weeks(40). Early vaccine effectiveness estimates for Omicron suggest there is significantly lower vaccine effectiveness from two doses of vaccine against symptomatic disease caused by Omicron variant compared to Delta variant. However from two weeks after a BNT162b2 booster dose, vaccine effectiveness against symptomatic disease caused by Omicron variant increases to 71% for those with a Vaxzevria primary course and 76% for those with a BNT162b2 primary course (26). The role of prior infection in combination with breakthrough infection following vaccination to protect against Omicron infection is not yet well defined.

As the number of individuals with previous SARS-CoV-2 infection increases, we will continue to see more reinfection cases and therefore it is vital that we continue to monitor the duration and nature of protection against reinfection in populations with high vaccine coverage and different emerging variants.

## Data Availability

The national datasets used in this research contain personal identifiable information which was used to link test results and are not available to the public. Aggregated positive test numbers are published daily on the UK coronavirus dashboard (https://coronavirus.data.gov.uk/) and UKHSA publishes data on numbers of possible reinfections in the National flu and COVID-19 surveillance reports (https://www.gov.uk/government/statistics/national-flu-and-covid-19-surveillance-reports-2021-to-2022-season). Any additional data requests should be made through the corresponding author.

