## Supplementary figure 1 for "Protective effect of a first SARS-CoV-2 infection from reinfection: a matched retrospective cohort study using PCR testing data in England"

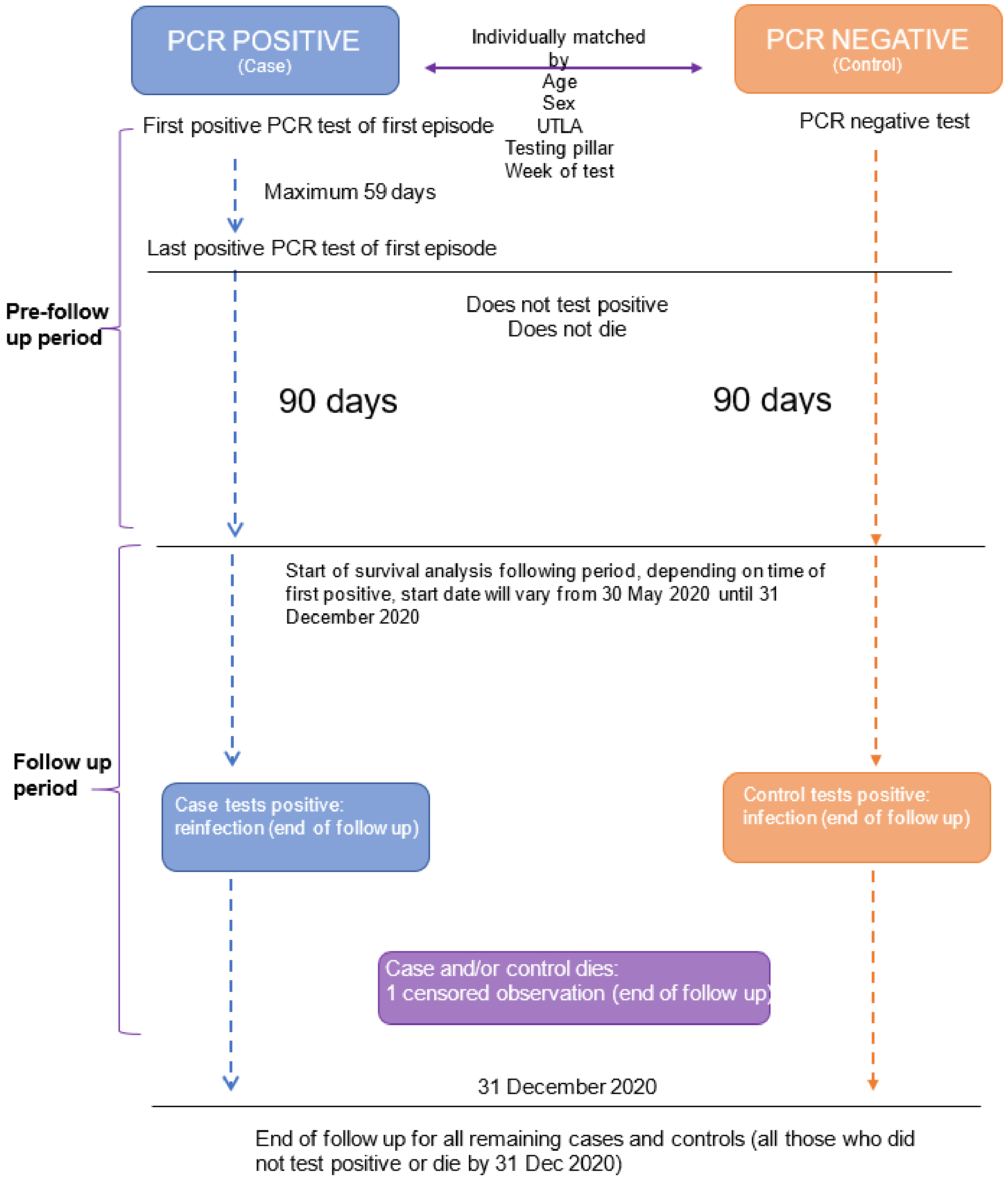

**Supplementary Figure S1** Diagram explaining the follow up time for PCR positive cases and their matched PCR negative control

**Supplementary table S1** Individuals that had symptoms at their second test within population that had a positive PCR test (post hoc population) during the follow up period and results of univariable and multivariable logistical regression

| | Number asymptomatic at positive test | Number with symptoms at positive test (% of total with symptom information) | Number with missing symptom information at second test (%) | Univariable odds ratio logistic regression for symptoms at second test | Multivariable odds ratio logistic regression for symptoms at second test * adjusted for all variables reported |
|---|---|---|---|---|---|
| **First test** | | | | | |
| Negative (Control) | 2,983 | 5860 (66.3) | 2875 (24.5) | 1 | 1 |
| Positive (Case) | 1,163 | 499 (30) | 599 (26.5) | 0.22 (0.19 0.24) | 0.23 (0.2 0.26) |
| **Sex** | | | | | |
| F | 3,075 | 3948 (56.2) | 2256 (24.3) | 1 | 1 |
| M | 1,071 | 2411 (69.2) | 1218 (25.9) | 1.75 (1.61 1.91) | 1.67 (1.51 1.84) |
| **Age** | | | | | |
| Aged 10-49 years | 2,005 | 4728 (70.2) | 1482 (18) | 1 | 1 |
| Aged 50 and above | 2,141 | 1631 (43.2) | 1992 (34.6) | 0.32 (0.3 0.35) | 0.33 (0.3 0.36) |
| **Month of first test** | | | | | |
| March | 65 | 146 (69.2) | 260 (55.2) | 1.17 (0.86 1.6) | 2.14 (1.48 3.08) |
| April | 970 | 1687 (63.5) | 1566 (37.1) | 0.91 (0.79 1.04) | 1.32 (1.14 1.54) |
| May | 1,535 | 1715 (52.8) | 944 (22.5) | 0.58 (0.51 0.66) | 0.84 (0.73 0.97) |
| June | 542 | 747 (58) | 312 (19.5) | 0.72 (0.62 0.84) | 0.86 (0.72 1.02) |
| July | 254 | 536 (67.8) | 150 (16) | 1.1 (0.92 1.32) | 1.19 (0.97 1.47) |
| August | 260 | 531 (67.1) | 99 (11.1) | 1.07 (0.89 1.28) | 1.2 (0.98 1.48) |
| September | 520 | 997 (65.7) | 143 (8.6) | 1 | 1 |
| **Region** | | | | | |
| London | 313 | 803 (72) | 427 (27.7) | 1 | 1 |
| East Midlands | 529 | 681 (56.3) | 275 (18.5) | 0.5 (0.42 0.6) | 0.56 (0.46 0.69) |
| East of England | 369 | 490 (57) | 351 (29) | 0.52 (0.43 0.62) | 0.62 (0.5 0.77) |
| North East | 253 | 372 (59.5) | 360 (36.5) | 0.57 (0.47 0.7) | 0.69 (0.54 0.87) |
| North West | 805 | 1509 (65.2) | 754 (24.6) | 0.73 (0.63 0.85) | 0.79 (0.65 0.94) |
| South East | 583 | 640 (52.3) | 496 (28.9) | 0.43 (0.36 0.51) | 0.47 (0.38 0.57) |
| South West | 204 | 190 (48.2) | 142 (26.5) | 0.36 (0.29 0.46) | 0.43 (0.33 0.57) |
| West Midlands | 514 | 673 (56.7) | 376 (24.1) | 0.51 (0.43 0.61) | 0.56 (0.46 0.69) |
| Yorkshire and Humber | 576 | 1001 (63.5) | 293 (15.7) | 0.68 (0.57 0.8) | 0.72 (0.59 0.87) |

| | | | | | |
|---|---|---|---|---|---|
| **Ethnicity** | | | | | |
| White | 3,210 | 4548 (58.6) | 2560 (24.8) | 1 | 1 |
| Asian or Asian British | 305 | 917 (75) | 332 (21.4) | 2.12 (1.85 2.43) | 1.56 (1.34 1.81) |
| Black or Black British | 173 | 152 (46.8) | 110 (25.3) | 0.62 (0.5 0.77) | 0.44 (0.34 0.56) |
| Mixed | 57 | 103 (64.4) | 40 (20) | 1.28 (0.92 1.77) | 0.97 (0.68 1.39) |
| Other ethnic groups | 81 | 169 (67.6) | 91 (26.7) | 1.47 (1.13 1.93) | 0.97 (0.72 1.31) |
| *Missing* | 320 | 470 (59.5) | 341 (30.2) | | |
| **IMD quintiles** | | | | | |
| 1 | 1,171 | 1780 (60.3) | 915 (23.7) | 1 | |
| 2 | 950 | 1419 (59.9) | 789 (25) | 0.98 (0.88 1.1) | |
| 3 | 782 | 1161 (59.8) | 660 (25.4) | 0.98 (0.87 1.1) | |
| 4 | 693 | 1087 (61.1) | 623 (25.9) | 1.03 (0.91 1.16) | |
| 5 | 539 | 901 (62.6) | 481 (25) | 1.1 (0.97 1.25) | |
| *Missing* | 11 | 11 (50) | 6 (21.4) | | |
| **First test route** | | | | | |
| Hospital testing | 1,111 | 1760 (61.3) | 2686 (48.3) | 1 | |
| Community testing | 3,035 | 4599 (60.2) | 788 (9.4) | 0.96 (0.88 1.04) | |

**Supplementary table S2** Individuals that had S gene target failure (SGTF) at their second test within population that had a positive PCR test from 15 November onwards (N=3128 with S gene target information, 5307 missing S gene target information) and results of univariable and multivariable logistical regression

| | Number positive for S gene at second test | Number with S gene target failure (% of total with S gene target) | Number tests after 15 Nov 2020 missing S gene target information (%) | Univariable odds ratio logistic regression for S gene negative | Multivariable odds ratio logistic regression for S gene negative at second test * adjusted for all variables reported |
|---|---|---|---|---|---|
| **First test** | | | | | |
| Negative (Control) | 1,632 | 1205 (42.5) | 4358 (60.6) | 1 | 1 |
| Positive (Case) | 132 | 159 (54.6) | 949 (76.5) | 1.63 (1.28 2.08) | 1.9 (1.43 2.51) |
| **Region** | | | | | |
| London, South East and East of England | 247 | 689 (73.6) | 2541 (73.1) | 1 | 1 |
| Rest of England | 1,517 | 675 (30.8) | 2766 (55.8) | 0.16 (0.13 0.19) | 0.14 (0.11 0.17) |
| **Age** | | | | | |
| Aged 10-49 years | 1,256 | 1027 (45) | 2823 (55.3) | 1 | 1 |
| Aged 50 and above | 508 | 337 (39.9) | 2484 (74.6) | 0.81 (0.69 0.95) | 0.9 (0.74 1.08) |
| **Month of first test** | | | | | |
| March | 34 | 30 (46.9) | 222 (77.6) | 0.64 (0.38 1.06) | 0.39 (0.22 0.71) |
| April | 377 | 307 (44.9) | 1591 (69.9) | 0.59 (0.48 0.72) | 0.4 (0.31 0.51) |
| May | 448 | 241 (35) | 1400 (67) | 0.39 (0.32 0.48) | 0.31 (0.24 0.39) |
| June | 188 | 76 (28.8) | 492 (65.1) | 0.29 (0.22 0.4) | 0.23 (0.16 0.32) |
| July | 149 | 98 (39.7) | 313 (55.9) | 0.48 (0.36 0.64) | 0.41 (0.29 0.57) |
| August | 238 | 156 (39.6) | 415 (51.3) | 0.47 (0.37 0.61) | 0.37 (0.28 0.49) |
| September | 330 | 456 (58) | 874 (52.7) | 1 | 1 |
| **Ethnicity** | | | | | |
| White | 1,258 | 953 (43.1) | 3926 (64) | 1 | 1 |
| Asian or Asian British | 283 | 195 (40.8) | 515 (51.9) | 0.91 (0.74 1.11) | 0.76 (0.61 0.96) |
| Black or Black British | 38 | 48 (55.8) | 208 (70.7) | 1.67 (1.08 2.57) | 1.05 (0.65 1.72) |

| | | | | | |
|---|---|---|---|---|---|
| Mixed | 17 | 24 (58.5) | 81 (66.4) | 1.86 (1 3.49) | 1.21 (0.59 2.48) |
| Other ethnic groups | 33 | 46 (58.2) | 155 (66.2) | 1.84 (1.17 2.9) | 0.9 (0.53 1.51) |
| *Missing* | 135 | 98 (42.1) | 422 (64.4) | | |
| **First test route** | | | | | |
| Hospital testing | 424 | 350 (45.2) | 2431 (75.9) | 1 | |
| Community testing | 1,340 | 1014 (43.1) | 2876 (55) | 0.92 (0.78 1.08) | |
| **Sex** | | | | | |
| F | 1,143 | 835 (42.2) | 3565 (64.3) | 1 | |
| M | 621 | 529 (46) | 1742 (60.2) | 1.17 (1.01 1.35) | |
| **IMD quintiles** | | | | | |
| 1 | 564 | 347 (38.1) | 1235 (57.5) | 1 | |
| 2 | 394 | 312 (44.2) | 1197 (62.9) | 1.29 (1.05 1.57) | |
| 3 | 318 | 284 (47.2) | 1092 (64.5) | 1.45 (1.18 1.79) | |
| 4 | 260 | 229 (46.8) | 988 (66.9) | 1.43 (1.15 1.79) | |
| 5 | 226 | 189 (45.5) | 781 (65.3) | 1.36 (1.07 1.72) | |
| *Missing* | 2 | 3 (60) | 14 (73.7) | | |
